# Association of smoking using with ALS risk and prognosis in China

**DOI:** 10.1101/2022.12.20.22283721

**Authors:** Wenchao Liu, XiaoGang Li, JingSi Jiang, Shuai Deng, Ying Song, Na Liu, Min Deng

## Abstract

**Objectives:** Cigarette smoking using have been posited as possible risk factors for amyotrophic lateral sclerosis (ALS), but few epidemiological studies supporting this hypothesis in China. We therefore explored the association between smoking with ALS incidence and prognosis in Chinese patients.

**Design:** Population-based case-control study.

**Setting, participants:** We performed a population-based case-control study in 812 ALS patients and 1500 matched controls. All the objects were recruited from Peking University Third Hospital, from January 2011 to December 2018 throughout China.

**Outcome measures:** Demographic data and information about premorbid cigarette smoking habits were collected at the time of diagnosis. The association of smoking with ALS was investigated using logistic regression analysis. Kaplan-Meier curves were used to compare survival time. Cox proportional hazards function and the hazard ratio were used to identify adjusted prognostic predictors.

**Results:** Current smokers had an increased risk of ALS (odds ratio = 1.66, 95% confidence interval (CI): 1.46, 1.87) compared to never smokers. Current cigarette pack years ≥20 had a significantly shorter median survival (63.89 months, IQR 55.90–71.87) compared with current cigarette pack years<20 (81.09 months, IQR 77.35–84.84) (p<0.001). Smoking habits were retained in Cox multivariable model.

**Conclusions:** Our study has proved current smoking is associated with an increased risk of ALS. Current cigarette pack years ≥20 is an independent negative prognostic factor for survival, with a dose–response gradient. Its influence is irrelevant to the presence ofchronic obstructive pulmonary disease (COPD) or to respiratory status at time of diagnosis. These results can be helpful for understanding and preventing ALS.

## Introduction

Amyotrophic lateral sclerosis (ALS) is a fatal neurodegenerative disorder of upper and lower motor neurons in the brain and spinal cord^1^. Death is usually from respiratory failure. In the absence of effective treatment, most patients die of respiratory failure within 2-5 years after onset. Only 5% -10% of patients survived beyond 10 years^2^. About 35 to 45% of patients with ALS have progressive cognitive abnormalities and 14% of ALS patients are diagnosed with frontotemporal dementia (FTD) ^3^. In most cases ALS appears sporadically in the population. Only about 10% of ALS cases are familial. In Europe and America, there are 1 or 2 new cases of ALS per year per 100,000 people. The total number of cases is approximately 3 to 5 per 100,000^2 4^, and is higher in men than in women, leading to a male-to-female ratio of 1.2–1.5^5 6^. Although its etiology remains unknown, several factors have been found to influence ALS phenotype, including the genetic factors, age, gender, premorbid diseases, life habits and physical activity. However, these studies and results are sparse and contradictory. Cigarette smoking, in particular, has attracted interest as a risk factor for ALS. In the worldwide, a number of epidemiologic studies have assessed the association of cigarette smoking with ALS incidence, but their results have been inconsistent^7-9^. The aim of our study was to assess the association of smoking with ALS incidence and outcome in a population-based case-control study of Chinese patients.

## Methods

### Patients, controls and information collected

A total of 812 ALS patients were recruited from Peking University Third Hospital between January 2011 and December 2018 throughout China, except Tibet, Taiwan, Hong Kong and Macao. All ALS patients were diagnosed by senior experts according to the El Escorial revised criteria and conformed to the criteria for probable and laboratory supported, possible or definite ALS ^10^. Patients with juvenile ALS, progressive muscular atrophy (PMA), progressive bulbar paralysis (PBP), and primary lateral sclerosis (PLS) diagnoses were excluded. Patients with additional diagnoses of other neurodegenerative diseases including essential tremor (ET), Parkinson’s disease (PD), Kennedy’s disease (KD) and multiple system atrophy (MSA) were excluded. And patients with interventions such as drugs and nasal intermittent positive pressure ventilation (NIPPV) were also excluded. One thousand five hundred age and gender matched controls were recruited from above three hospitals at the same time. The eligibility criteria of controls included: no diagnosis of ALS or other neurodegenerative disease, and no family history of ALS. Survival time was defined as the interval time between date of onset and date of death, tracheotomy and follow-up date. Severity of ALS was assessed by the ALSFRS-R scale ^11^. Rate of disease progression (the decline rate for ALSFRS-R score) was calculated as the mean monthly number of point loss from the time of diagnosis to telephone follow-up, calculated in months. Information on the demographic data and clinical data were recorded, including age of onset, gender, dyspnea, breathing machine, marriage status, nationality, site of onset (bulbar or limb), body mass index (BMI), diagnostic delay (duration from onset to diagnosis), El Escorial classification at diagnosis, ALSFRS-R score at diagnosis and follow-up months. And for each patient we collected information about their cigarette smoking habits and chronic obstructive pulmonary disease (COPD). The cigarette means volume of cigarettes, not e-cigarettes. Patients’ smoking status was defined as current smokers, former smokers and never smokers. Smoking intensity was defined as current cigarette pack years<20 (former smokers and never smokers were also included) (The pack-years of smoking were calculated by multiplying the average number of packs of cigarettes smoked per day by the number of years the person had smoked) and current cigarette pack years ≥20^12^. COPD was classified according to the Global Initiative for Chronic Obstructive Lung Disease (GOLD) guidelines^13^. Forced vital capacity (FVC) per cent of predicted and forced expiratory volume in the 1 s (FEV1) were performed and annotated. FEV1/FVC ratio was calculated for diagnosing the severity of COPD. All subjects provided written informed consent to participate in the study in accordance with the process approved by the Institutional Ethics Committee of Peking University Third Hospital.

## Statistical methods

Comparisons between means were made with Student’s t-test or analysis of variance (ANOVA); comparison between categorical variables was made with χ2 test. All tests were two-tailed. Levene’s test was used to confirm the equality of variances. Non-normal distribution was assessed with the Kruskal-Wallis test. The associations between smoking and risk of ALS were evaluated by means of univariate analysis using logistic regression and multivariate logistic regression. Survival was calculated from onset to death, tracheostomy or follow-up date, using the Kaplan-Meier method, and compared with the log-rank test; when more than two ordinal strata were assessed, the linear trend for factor level test was used. No patients were lost to follow-up.

Multivariable analysis was performed with the Cox proportional hazards model. A p level <0.05 was considered significant. Statistical analyses were carried out using the SPSS V.21.0 statistical package (SPSS, Chicago, Illinois, USA).

## Results

Although the patients were conducted at Peking University Third Hospital, they came from throughout China. During the study period, we identified 812 ALS cases and selected 1500 matched controls. All 812 ALS patients were included in the case group, including 531 male patients (65.39%) and 281 female patients (34.61%). The male to female ratio for the cohort was 1.89:1. The mean age at onset was 49.49±11.16 years; the mean age of male (49.83±10.86) was similar to female (48.84±11.70) (p=0.232); the mean age of patients with breathing machine (53.18±11.89) was significantly older than that of patients without breathing machine (49.30±11.10) of patients (p=0.034); the mean age at bulbar-onset (52.11±11.41) was significantly older than mean age at limb-onset (48.97±11.05) of patients (p=0.003). The demographic and clinical data of case group and control group, as well as of their premorbid smoking status and COPD were shown in Table 1.

**Table 1.**
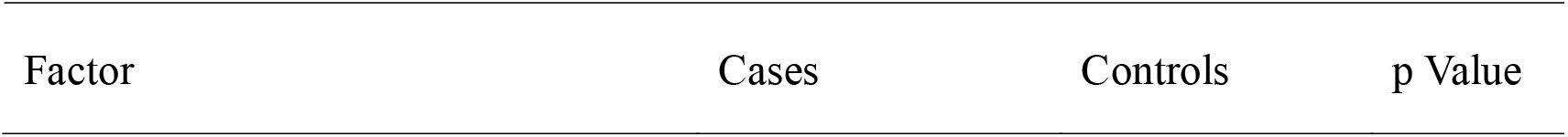
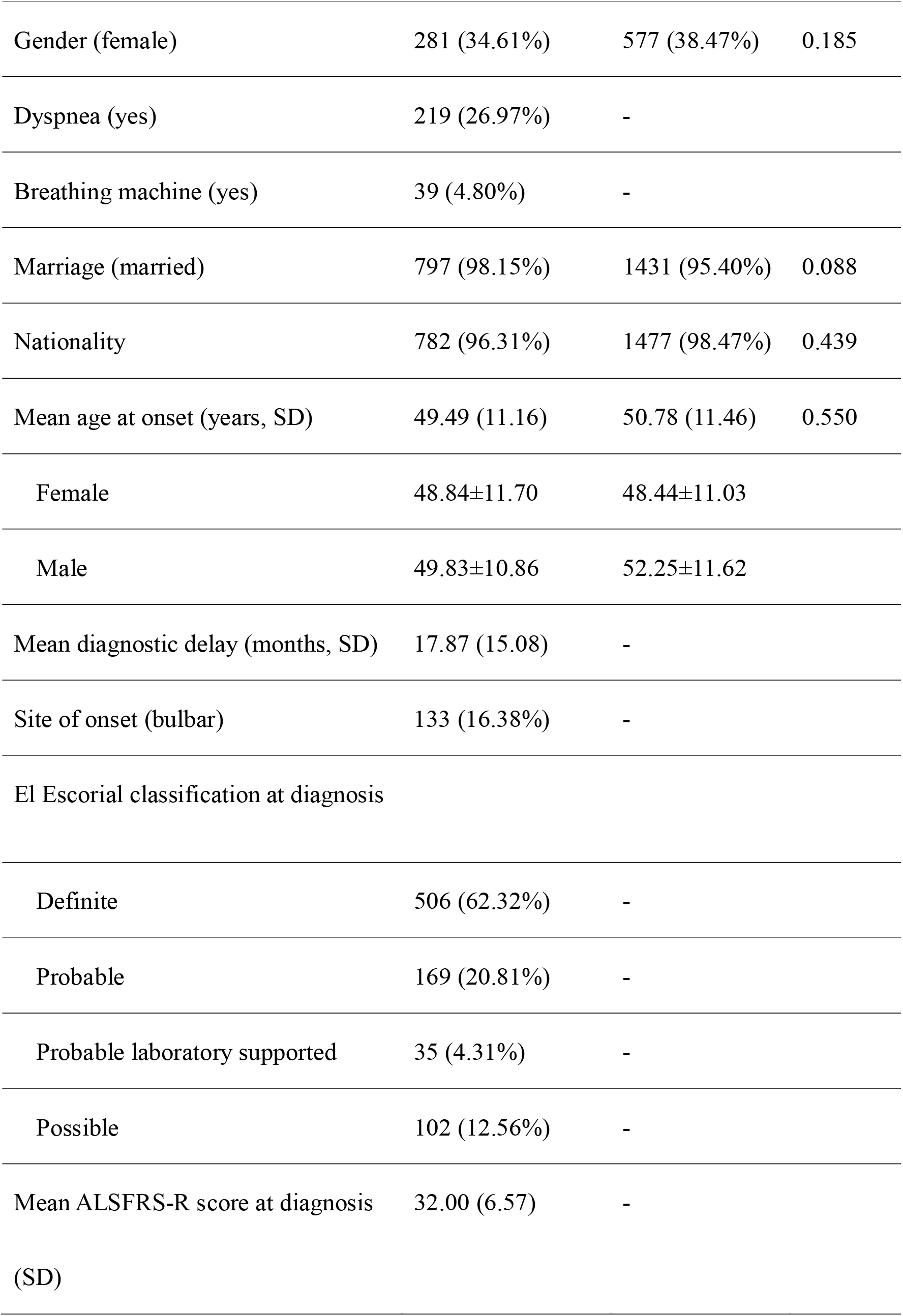
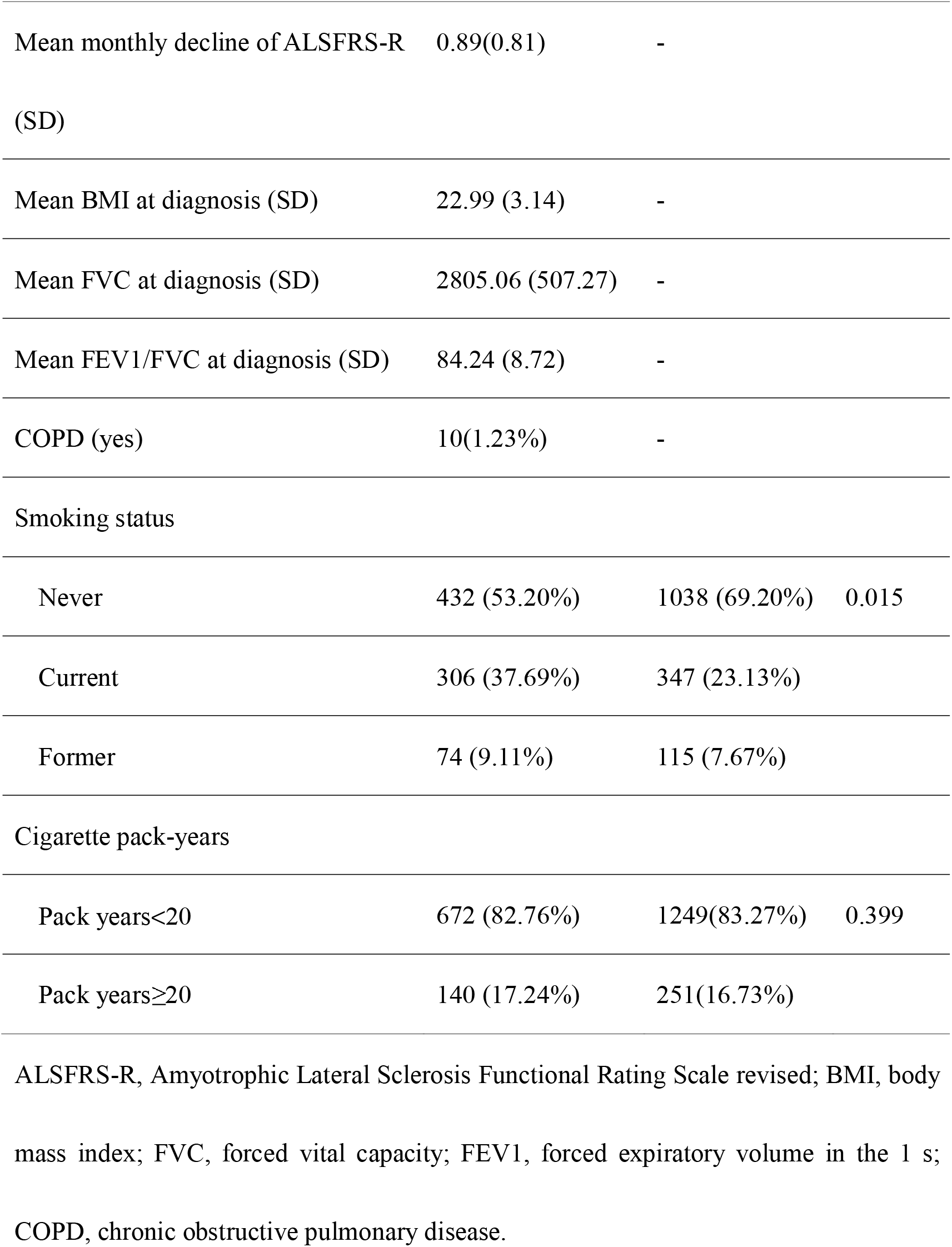
Demographic and clinical characteristics of all subjects.

In our study, the number of current smokers in case group were higher than control group (p=0.015). Factors with p value less than 0.1 were included in the multivariate analysis. Multivariate analyses in ALS patients showed an increased risk of current smokers (odds ratio = 1.66, 95% confidence interval (CI): 1.46, 1.87) compared with never smokers (p=0.003).

In case group of our study, 672 patients (82.76%) were current smokers with <20 pack years at the time of ALS onset, and 140 (17.24%) were current smokers with ≥20 pack years. ≥20 pack-year current smokers were more likely to be male (pack years ≥20, 82.86%; pack years<20, 61.76%; p<0.001), use breathing machine (pack years ≥20, 9.29%; pack years<20, 3.87%; p=0.009) and have an older age at onset (pack years ≥20, 56.68 years(SD 8.51); pack years<20, 47.99 years (SD 11.07); p<0.001). A total of 10 patients (pack years ≥20, 1.43%; pack years<20, 1.19%) were affected by COPD at the time of ALS symptom onset. Patients with COPD had a similar age at onset to patients without COPD (53.40 years (SD 12.49) vs 49.44 years (SD 11.14); p=0.265). The median survival time of patients with COPD was similar to patients without COPD (COPD, 52.80 months, IQR 27.97–77.63; non-COPD, 78.30 months, IQR 74.79–81.80) (p=0.271).

Kaplan-Meier analysis revealed that no significant differences were found in patients with different smoking status. However, smoking intensity was correlated with survival time. Current smokers with pack years ≥20 had a significantly shorter median survival (63.89 months, IQR 55.90–71.87) than those with pack years <20 (81.09 months, IQR 77.35–84.84) (p<0.001) (Figure 1). This difference was present in men and women and was not modified stratifying by age at onset, site of onset, and so on. Stratifying by FVC and FEV1/FVC ratio, the negative effect of smoke on survival was still present (data not shown). In addition, stratification for COPD did not modify the effect of premorbid smoking habits on survival (data not shown). Current cigarette pack years ≥20 was significantly correlated to the mean monthly decline of ALSFRS-R (p=0.002), and it was not correlated with FVC, FEV1/FVC ratio, body mass index (BMI) at diagnosis (p=0.955; p=0.723; p=0.154). We also found dyspnea was significantly correlated to the breathing machine, FVC, FEV1/FVC ratio (p=0.007; p=<0.001; p<0.001).

**Fig. 1.**
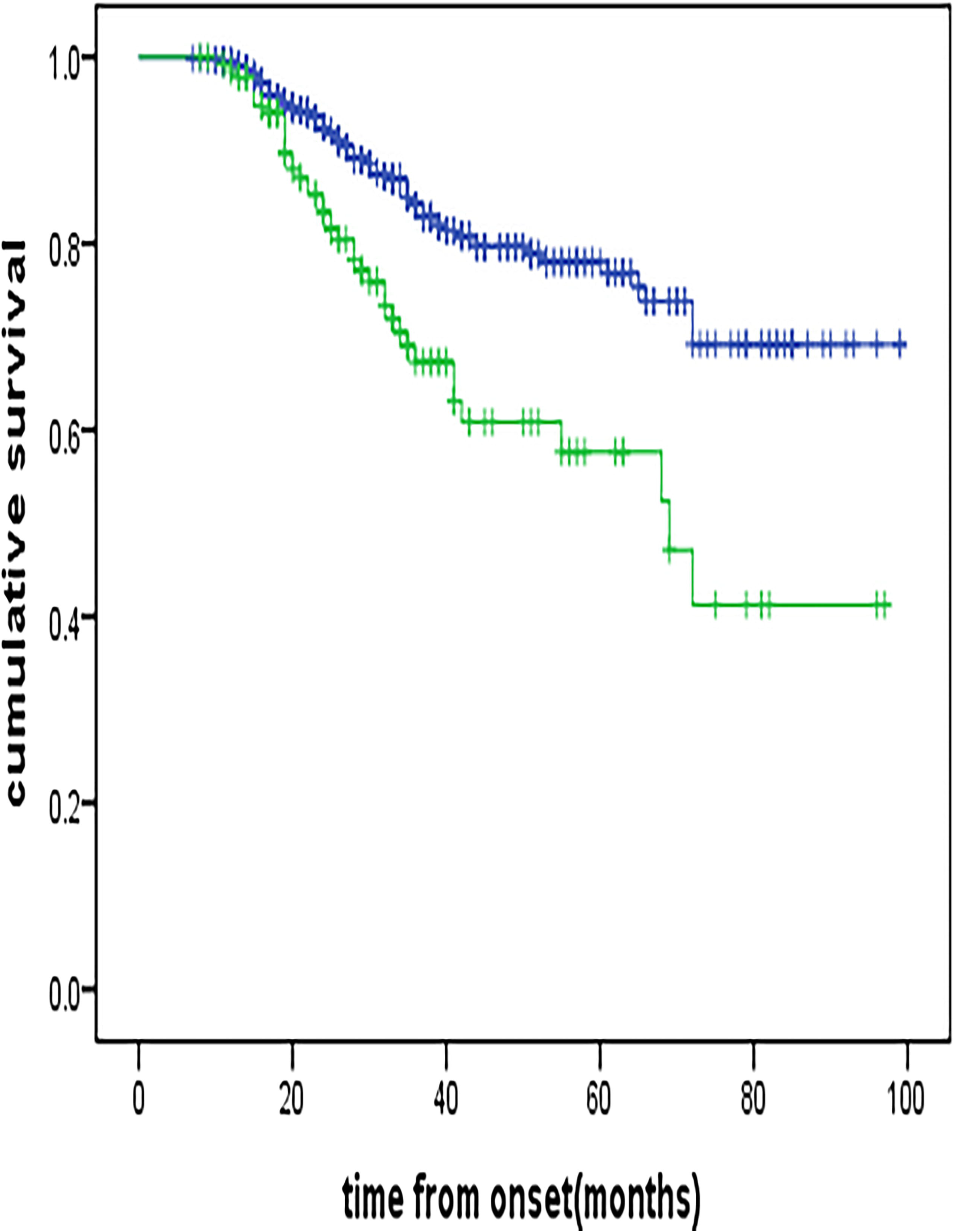
Kaplan-Meier curves by smoking intensity at time of amyotrophic lateral sclerosis onset. The blue line represents current cigarette pack years<20, the green line current cigarette pack years_≥_20; p=0.000, linear trend.

Therefore, we performed a multivariate analysis. The parameters included age of onset, gender, site of onset, diagnostic delay, ALSFRS-R decline (points/month). Cox model confirmed that current cigarette pack years ≥20 was an independent negative prognostic factor with an increased HR for currently cigarette pack years <20 of 1.75 (95% CI 1.15 to 2.67, p=0.009). COPD is not an independent negative prognostic factor. The main significative model was shown in Table 2.

**Table 2.** Multivariable model including all cases.

| Factor | HR (95% CI) | p Value |
| --- | --- | --- |
| Age (years) |  |  |
| 18–49 | 1 |  |
| 50–59 | 1.69 (1.04 to 2.74) | 0.035 |
| 60–69 | 2.62 (1.56 to 4.42) | 0.000 |
| 70+ | 2.30 (0.94 to 5.63) | 0.068 |
| Site of onset |  |  |
| Bulbar | 1 |  |
| Spinal | 0.90 (0.53 to 1.50) | 0.672 |
| Diagnostic delay |  |  |
| >1 year | 1 |  |
| ≤1 year | 3.27 (2.11 to 5.06) | 0.000 |
| Smoking intensity |  |  |
| Current cigarette pack years<20 | 1 |  |
| Current cigarette pack years≥20 | 1.75 (1.15 to 2.67) | 0.009 |
| ALSFRS-R decline (points/month) |  |  |
| <0.75 | 1 |  |
| ≥0.75 | 3.00 (1.95 to 4.64) | 0.000 |
| Variables included in the model: age at onset (18–49, 50–59, 60–69, 70+); site of onset (bulbar; spinal); ALSFRS-R decline (<0.75, ≥0.75 points/months); body mass index at diagnosis (underweight <18.5; normal weight, 18.5–24.99; overweight, 25–29.99; obese classes I, II and III, ≥30 kg/m<sup>2</sup>); diagnostic delay (>1; ≤1 year), smoking intensity (pack years<20, pack years≥20). ALS, amyotrophic lateral sclerosis; ALSFRS-R, ALS Functional Rating Scale revised. |  |  |

Variables included in the model: age at onset (18–49, 50–59, 60–69, 70+); site of onset (bulbar; spinal); ALSFRS-R decline (<0.75, ≥0.75 points/months); body mass index at diagnosis (underweight <18.5; normal weight, 18.5–24.99; overweight, 25–29.99; obese classes I, II and II, ≥30 kg/m2); diagnostic delay (>1; ≥1 year), smoking intensity (pack years<20, pack years ≥20). ALS, amyotrophic lateral sclerosis; ALSFRS-R, ALS Functional Rating Scale revised.

## Discussion

There are few studies on ALS in the Chinese population. Most ALS researches were based on epidemiological and genetic studies of ALS patients from Caucasian populations of European origin. However, the Chinese are significantly different from Caucasians in terms of ethnic, social, cultural background and quality of healthcare^14 15^. Therefore, the epidemiological characteristics of ALS in Chinese are different from those in other countries. The mean age of onset in our study is 49.49 years old (SD 11.16), younger than patients form Japan and Europe, 61.0 ^16^and 63.79 years old^17^, respectively. However, in India, it is 51.6 years old^18^, lower than that in most other regions around the world. It has been suggested that age of onset is younger in less developed regions and higher level of environmental pollution areas. The male to female ratio for the cohort was 1.89:1, while in Europe it was 1.2:1-1.5:1. There were more than four-fifth of ALS patients (83.62%) to have limb onset in the study (Table 1), higher than 61.7% limb onset in Europe^19^, 59.9% limb onset in Italy^20^, and 72.9% limb onset in India^21^. Even patients onset from the limb different carried mutated genes (SOD1, TARDBP, FUS/TLS, and C9ORF72) in Chinese with familial ALS ^5^. The diagnostic delay of ALS in our study (17.87 months) is longer than Europe and the United States (12 months)^1^, possibly due to differences in health care systems or the seemingly more aggressive disease phenotype in Europe and the United States. The overall male-to female ratio, site of onset, diagnostic duration in this study was different from those in our previous study about mutated genes associated with familial ALS^5^.

We have explored the effect of smoking on ALS phenotype and outcome in a population-based cohort of Chinese patients. Current cigarette pack years ≥20 is a strong independent modifier of prognosis, with a decreasing gradient, current cigarette pack years ≥20(63.89 months, IQR 55.90–71.87) having a reduction of overall survival of 17.2 months compared with current cigarette pack years <20(81.09 months, IQR 77.35–84.84). Most published studies indicate that it increases the risk of developing ALS, and others indicate that smoking was a predictor of ALS in women, and of mortality in female ALS patients, but not in men^22-24^. However, few studies have assessed its influence on ALS outcome, with some inconsistencies. In our study, current cigarette pack years ≥20 displayed one effect on ALS outcome: patients with currently cigarette pack years ≥20 at disease onset had a shorter survival, different from the study of Andrea Calvo et al (current smokers had a significantly shorter median survival compared with former and never smokers; also, COPD adversely influenced patients’ prognosis)^24^, also, different from the study of Cui et al (the risk factors of ALS included smoking,but years and numbers of smoking did not show correlation with ALS in Chinese people)^25^. The study of Al-Chalabi et al used Mendelian randomization methods to assess the relationship between smoking and ALS and found that smoking didn’t causes ALS^26^. However, in another paper they revealed that there was weak evidence of a positive effect of current smoking on the risk of ALS^27^. The effect of smoking on survival was independent from other prognostic factors, including age, gender, site of onset, diagnostic delay, El Escorial classification at diagnosis, COPD and respiratory function as measured by FVC and FEV1/FVC ratio. Another paper of Peters S et al further confirmed that the correlation between smoking and ALS. They found smoking duration rather than intensity is more relevant before adjustment for time-since-quitting and high-intensity smoking with a short duration is more damage than low-intensity smoking with a long duration^28^.

Cigarette smoking could increase the risk of developing ALS through several mechanisms, including inflammation, oxidative stress, nitric oxide and neurotoxicity caused by heavy metals and other chemical compounds present in cigarette smoke. Oxidative stress is known to be a major contributor for developing degenerative diseases. Studies have shown the potential of oxidative burden in worsening the cellular degeneration. Certain molecules such as HNE, MDA, lsoprostanes and TBARS are well studied as biomarkers for increase of oxidative stress through LPO. A study found that cigarette smoking may enhance LPO levels^29^. Smoking could also act at epigenetic level, through an aberrant methylation of DNA^30^. COPD has not been found to be an independent negative prognostic factor in ALS. The prevalence of COPD in China is less than recent epidemiological studies in the Italian population^31 32^, and our study is significantly less than study in Chinese population. The prevalence of COPD is especially common in people over 60 aged, but in our study, the mean age at onset in patients with ALS is 49.49, only 125 patients (15.39%) over 60 aged, which can be an explanation. In addition, the small number of samples may also lead to the small quantity of patients with COPD. Therefore, our study did not find COPD influenced patients’ prognosis.

Several limitations of the present study should be noted. First, we excluded other spectra of possible ALS populations. Second, sample size is very important, and a small sample size can result in insufficient power, thus bringing about type 1 error in the results. Last, other factors may also be implicated as prognostic predictors, such as cognitive status, which we did not investigate at the time of the study.

## Conclusions

Our population-based case-control study found that the age at onset of ALS in China was earlier than other countries. Current smoking is associated with an increased risk of ALS. Cigarette smoking (pack years ≥20) is an independent negative prognostic factor for survival, with a dose–response gradient, independent from age, gender and other known factors, including respiratory function and COPD. According to these findings, neurologists should consider to recommend to their patients with ALS the cessation of smoking as a measure to significantly improve their outcome. This study indicates that environmental factors and personal habits represent risk factors for ALS onset and can also influence its phenotype and prognosis. These results can be helpful for understanding and preventing ALS.

## Data Availability

Please include a statement regarding the availability of all data referred to in the manuscript.
Example statements:
All data produced in the present study are available upon reasonable request to the authors.
All data produced in the present work are contained in the manuscript.
All data produced are available online at.

## Acknowledgements

We thank all participants who took part in this project. Written informed consent for publication of the patients’ clinical information and molecular data were obtained from their families.

## Contributors

DM was responsible for the study concept and design. LWC, LXG JJS, DS, SY, and LN did the data and project management. LWC and DS did the data cleaning and analysis. LWC, LXG, and JJS interpreted the data and drafted the manuscript. DM is responsible for the overall content as the guarantor. All the authors approved the final manuscript as submitted and agree to be accountable for all aspects of the work.

## Data availability statement

The data that support the findings of this study are available from the authors upon reasonable request and with permission of Peking University Third Hospital.

### Patient and public involvement

Patient and public involvement Patients and/or the public were involved in the design, or conduct, or reporting, or dissemination plans of this research. Refer to the Methods section for further details.

### Ethics approval and consent to participate

The Ethical Committee of Peking University Third Hospital has approved this research, and all the participants consented to this study with written informed consent.

